# Using the Association Between Antibiotic Susceptibility and Genetic Relatedness to Rescue Old Drugs for Empiric Use

**DOI:** 10.1101/19006106

**Authors:** Derek R MacFadden, Bryan Coburn, Karel Břinda, Antoine Corbeil, Nick Daneman, David Fisman, Robyn S Lee, Marc Lipsitch, Allison McGeer, Roberto G. Melano, Samira Mubareka, William P Hanage

## Abstract

**Background:** Rising rates of antibiotic resistance have led to the use of broader spectrum antibiotics and increasingly compromise empiric therapy. Knowing the antibiotic susceptibility of a pathogen’s close genetic relative(s) may improve empiric antibiotic selection.

**Methods:** Using genomic and phenotypic data from three separate clinically-derived databases of *Escherichia coli* isolates, we evaluated multiple genomic methods and statistical models for predicting antibiotic susceptibility, focusing on potentially rapidly available information such as lineage or genetic distance from archived isolates. We applied these methods to derive and validate prediction of antibiotic susceptibility to common antibiotics.

**Results:** We evaluated 968 separate episodes of suspected and confirmed infection with *Escherichia coli* from three geographically and temporally separated databases in Ontario, Canada, from 2010-2018. The most common sequence type (ST) was ST131 (30%). Antibiotic susceptibility to ciprofloxacin and trimethoprim-sulfamethoxazole were lowest (<=72%). Across all approaches, model performance (AUC) ranges for predicting antibiotic susceptibility were greatest for ciprofloxacin (0.76-0.97), and lowest for trimethoprim-sulfamethoxazole (0.51-0.80). When a model predicted a susceptible isolate, the resulting (post-test) probabilities of susceptibility were sufficient to warrant empiric therapy for most antibiotics (mean 92%). An approach combining multiple models could permit the use of narrower spectrum oral agents in 2 out of every 3 patients while maintaining high treatment adequacy (∼90%).

**Conclusions:** Methods based on genetic relatedness to archived samples in *E. coli* could be used to rescue older and typically unsuitable agents for use as empiric antibiotic therapy, as well as improve decisions to select newer broader spectrum agents.

**Summary:** Rapid genomic approaches that capitalize on the association between genetic relatedness and phenotype can improve our selection of antibiotics, allowing us to rescue older drugs for empiric use and better select newer and broader spectrum agents.

## Background

Antibiotic resistance is a major global threat to public health[1]. Antibiotic resistant organisms (AROs) and mechanisms of antibiotic resistance are selected through the use of antibiotics in humans, animals, and environments[2]. The prevalence of AROs in human infections has been increasing in many regions and across many different bacterial species[1,3]. As a result, empiric antibiotic therapy (administered prior to knowledge of the organism’s antibiotic susceptibility phenotype) has become increasingly challenging for both community- and hospital-acquired infectious syndromes. Inadequate empiric therapy, i.e., treatment that does not include an agent to which the etiologic pathogen is susceptible, has been associated with worse patient outcomes[4–6]. Moreover, increasing antibiotic resistance in common infections leads to more frequent use of broader spectrum antibiotic agents, with added toxicity and enhanced selection of antibiotic resistance by targeting a wider array of pathogenic, opportunistic, and commensal bacteria.

Reducing the time from presentation and sample collection to reporting of antibiotic susceptibility has long been touted as a potential means to improve early adequate therapy and reduce the use of unnecessarily broad antibiotic agents[7,8]. Development of rapid diagnostic tests that can narrow these ‘windows’ of empiric antibiotic therapy are the focus of active research, but translation of these tests to clinical practice has been slow, due to inconsistency between individual or combined genetic loci and expected phenotype, specialized equipment, cost, challenges of commercialization, and poor integration into clinical workflow[7]. Recently, genomic approaches have been identified as rapid diagnostic tests, offering the promise of culture-independent (and -dependent) identification of: (1) species; (2) relationship(s) to genetic neighbors/groups/clusters from databases of known isolates; and (3) prediction of antibiotic resistance based on (2). However, the traditional approach to predicting antibiotic resistance rests on identification of individual resistance loci to predict phenotype. This requires a high-quality database of resistance-causative elements and is further complicated by significant cost, large physical space requirements, complicated workflow, limited expertise, and long sequencing/bioinformatic processing times even with real-time sequencing technologies[9]. On the other hand, a recently introduced alternative approach called genomic neighbor typing infers antibiotic resistance and susceptibility by identifying sample’s closest relatives in a database of genomes with known phenotypes [10]. This relies on strong correlation between phylogenetic group and resistance phenotype, which is observed for many bacteria[11–13].

As neighbor typing uses all genomic data available from a given set of reads, identification of a best match isolate or lineage (i.e. a genetically related cluster or group such as multi-locus sequence type) can occur within minutes (as opposed to hours or days for loci-based approaches depending upon the sequencing technology used). Proof of principle has been demonstrated for *Streptococcus pneumoniae* and *Neisseria gonorrhoeae*, with determination of resistance or susceptibility within ten minutes of Oxford Nanopore Technologies’ © MinION sequencing of cultured isolates and respiratory metagenomic samples[10]. Limited data also suggest that the association between antibiotic susceptibility phenotype and lineage may also hold true for *Enterobacteriaceae[14,15]*, but this approach requires further validation. It is also unknown whether predicting antibiotic susceptibility phenotype based on the phenotype of the nearest genetic neighbor provides advantages over using the average phenotype of a broader (or higher level) lineage (e.g. ST, clonal complex, or cluster). In order to understand the potential clinical application of these techniques, we sought to validate the association between genetic relatedness (using nearest neighbor and lineage markers) and antibiotic susceptibility phenotype in the most common Gram-negative pathogen in humans, *Escherichia coli*.

## Methods

### Study Design

We performed a retrospective study to evaluate whether genetic relatedness can predict antibiotic susceptibility in *E. coli* isolates from 968 episodes of suspected and confirmed human infection. Three separate datasets were combined for this analysis and include: (Dataset 1) 411 *E*.*coli* isolates from bloodstream infections at Sunnybrook Health Sciences Centre (SHSC), a single tertiary care medical centre in Toronto, Canada, collected over the years 2010-2015; (Dataset 2) 177 *E*.*coli* from suspected urinary tract infections from SHSC for the year 2018; and (Dataset 3) 380 multi-drug resistant (MDR) *E*.*coli* isolates from urinary sources from the Canadian province of Ontario collected in 2010 and 2015, where MDR was defined as resistance to at least three different classes of routinely tested antibiotics.

### Resistance Phenotype

Antibiotic susceptibility phenotypes for ciprofloxacin (fluoroquinolones), trimethoprim-sulfamethoxazole (sulfonamides), ceftriaxone (3rd generation cephalosporins), gentamicin (aminoglycosides), and ertapenem (carbapenems), were determined for each isolate using Vitek 2 AST cards. Clinical laboratory standards institute (CLSI) 2015 breakpoints were employed for determining susceptible and non-susceptible phenotypes for all datasets (Supplementary Table 1). For Dataset 2, only formal ESBL testing was available (not ceftriaxone MICs reported) and as such we classified all non-ESBL producing *E.coli* as susceptible to ceftriaxone. Given that we calculated susceptibility using MIC’s and static breakpoints for all datasets, there were no temporal changes in the interpretation of susceptibility. We have considered all non-susceptible isolates as resistant throughout this paper.

### Whole Genome Sequencing

Genomes for each dataset were sequenced separately using a NextSeq High Output platform with Nextera Library Preparation with mean coverages of 134X, 90X, and 81X for dataset 1, 2, and 3 respectively. Further sequencing details can be found in the Supplementary Methods.

### Overall Prediction Approach

As overfitting could be a major limitation of our approach, and to simulate potential clinical implementation strategies, we externally validated previously collected derivation datasets (Dataset 1 and 3) for predicting the susceptibility of isolates from the most recent dataset (Dataset 2). Where applicable, we also evaluated the sets internally with bootstrapping to adjust for optimism (overfitting). We followed the general principles of the TRIPOD statement for reporting of multivariable prediction models [16].

Four prediction model approaches were employed and are described in detail in the Supplemental material. Briefly the first model is an *ST Parametric Model Approach* which uses ST (marker of lineage) as a categorical predictor within a logistic regression model to predict the probability of susceptibility. The second model is a *Cluster Parametric Model Approach* that uses labelled genetic clusters (marker of lineage) as categorical predictors in a logistic regression model to predict the probability of susceptibility. The third model is an *ST Reference Database Approach* that uses the average prevalence of susceptibility of an antibiotic for a given ST (marker of lineage) as the predicted probability of susceptibility. And the fourth model is a *Best Genetic Match Reference Database Approach* that uses the susceptibility phenotype of the best genetic match (nearest neighbor) in a reference database as the predicted susceptibility. The above analyses were performed separately for each antibiotic.

To further simulate the antibiotic decision-making process, we explored two different scenarios where an antibiotic was selected based upon sequential model outputs, either favouring narrow spectrum agents or favoring high likelihood of adequacy of coverage. We called these *Sequential Decision-making Models*. Further details on these methods are described in the Supplemental material. Research ethics board approval was obtained for this study at SHSC.

## Results

### Description of the Datasets

We collected and sequenced the genomes of 968 unique *E.coli* isolates from separate clinical episodes of suspected infection, across three datasets. The characteristics of each dataset are shown in Table 1 below.

**Table 1.** Characteristics of datasets.

|  | Dataset 1 | Dataset 2 | Dataset 3 |
| --- | --- | --- | --- |
| **Number of Isolates (n=968)** | 411 | 177 | 380 |
| **Collection Period** | 2010-2015 | 2018 | 2010 and 2015 |
| **Location** | Toronto (City) | Toronto (City) | Southeastern Ontario |
| **Location Type** | Hospital Lab | Hospital Lab | Hospital Lab |
| **Inpatient/Outpatient** | Inpatient | In/Outpatient | In/Outpatient |
| **Anatomic Site** | Blood | Urine | Variable |
| **Sampling Bias** | None | None | Multi-drug Resistant (MDR) |
| **Antibiotic Susceptibility\*** |  |  |  |
| *Ciprofloxacin* | 297 (72) | 120 (68) | 118 (31) |
| *Ceftriaxone* | 357 (87) | 155 (88) | 236 (62) |
| *Gentamicin* | 355 (86) | 156 (88) | 229 (60) |
| *Trimethoprim-Sulfamethoxazole* | 292 (71) | 125 (71) | 79 (21) |
| *Ertapenem* | 410 (99) | 177 (100) | 338 (89) |
| **Predominant ST\*** |  |  |  |
| **1193** | 14 (3.4) | 15 (8.5) | 21 (5.5) |
| **127** | 15 (3.6) | 7 (4.0) | 3 (0.8) |
| **131** | 87 (21) | 36 (20) | 170 (45) |
| **38** | 7 (1.7) | 2 (1.1) | 12 (3.2) |
| **405** | 9 (2.2) | 1 (0.6) | 11 (2.9) |
| **648** | 6 (1.5) | 8 (4.5) | 17 (4.5) |
| **69** | 22 (5.4) | 13 (7.3) | 23 (6.1) |
| **73** | 57 (14) | 20 (11) | 20 (5.3) |
| **95** | 58 (14) | 19 (11) | 12 (3.2) |
| **Other** | 136 (33) | 56 (32) | 91 (24) |

\*N(%)

### Genetic Relatedness of Different Datasets and Antibiotic Resistance Phenotype

To illustrate the linkage between relatedness and resistance, a genetic tree was constructed using Mash distances, associated ST, antibiotic susceptibility phenotype, and source dataset (Figure 1). Broad genetic clusters tended to match or be nested within STs, and closely related isolates have similar antibiograms.

**Figure 1.**
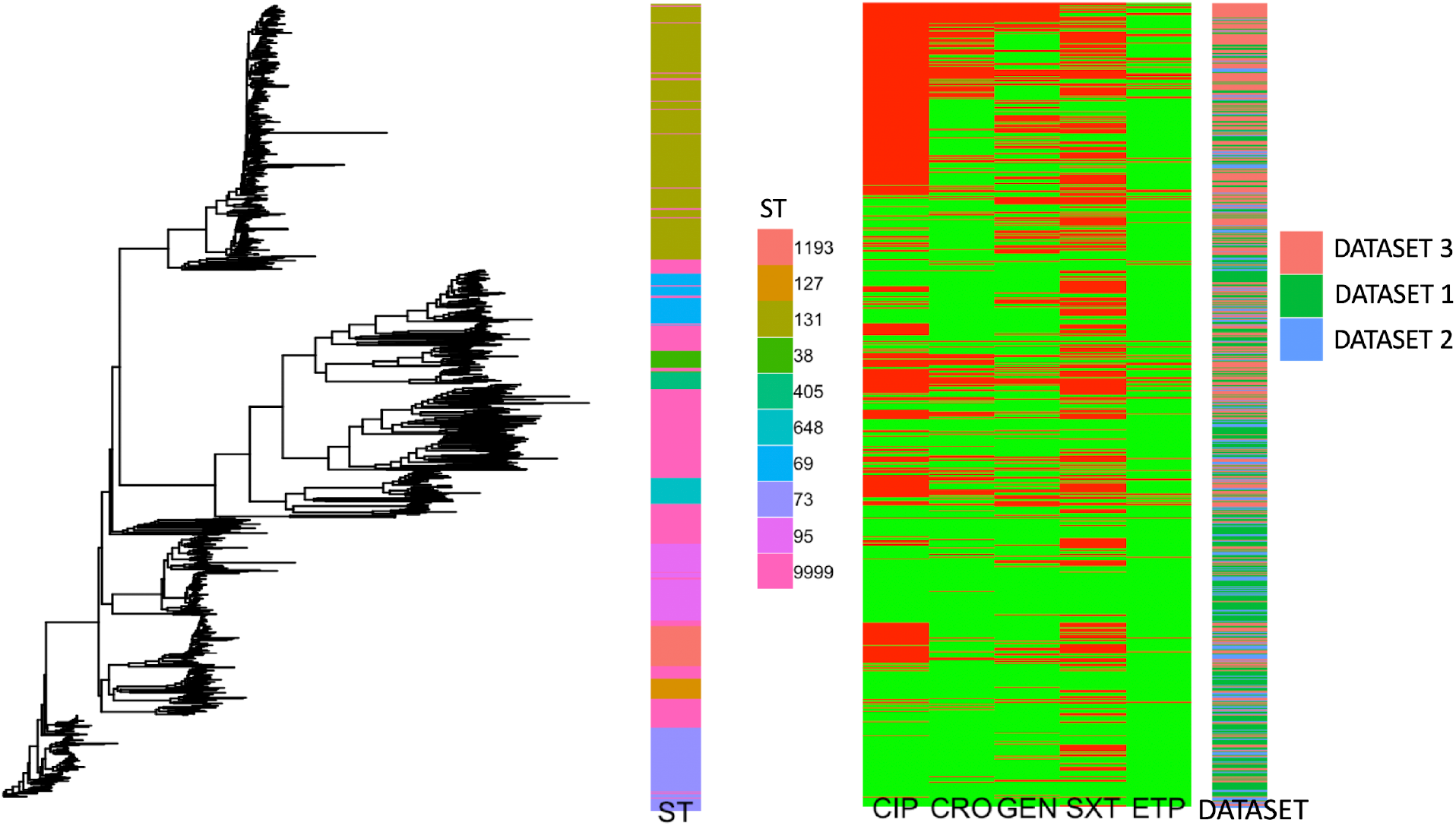
Mash tree (left), ST, phenotypic susceptibility by antibiotic, and dataset, by individual isolate. Antibiotic susceptibility is denoted in green and resistance in red, with antibiotics abbreviated as: CIP (ciprofloxacin), CRO (ceftriaxone), GEN (gentamicin), SXT (trimethoprim-sulfamethoxazole), ETP (ertapenem). ST 9999 represents all remaining or unknown STs.

From the Mash tree illustrating high level relationships between the genomes (Figure 1), there are clear genetic clusters that emerge. These phylogenetic groups are generally nested within specific STs, with ST131 being the most prevalent in each dataset (Table 1). Unsurprisingly, there is a higher prevalence of resistance for almost all antibiotic groups in the MDR dataset (Dataset 3) compared to Datasets 1 and 2. Despite the fact that the three datasets are separated on different scales (temporally, geographically, anatomically), they show genetic diversity that is well distributed across different phylogroups (Figure 1).

### ST Parametric Model Approach (Lineage-based)

We calculated the AUCs and test characteristics for predicting antibiotic susceptibility of Dataset 2 isolates using a parametric logistic regression model with STs as categorical predictors, across a variety of derivation datasets (Supplementary Table 3). AUCs ranged from 0.89-0.91 for ciprofloxacin, 0.77-0.80 for ceftriaxone, 0.68 to 0.75 for gentamicin, and 0.6-0.73 for trimethoprim-sulfamethoxazole. For ertapenem, we performed internal derivation for Dataset 1, Dataset 3, and Datasets 1 and 3 combined (Dataset 2 could not be used as there was 100% susceptibility to ertapenem), and found apparent and optimism adjusted AUCs ranging from 0.7- 0.99 and 0.67-0.99 respectively (Supplementary Table 2).

### Cluster Parametric Model Approach (Lineage-based)

We calculated the AUC and test characteristics for predicting antibiotic susceptibility (internally) for each dataset with a parametric logistic regression model with clusters as categorical predictors, using a variety of derivation datasets (Supplementary Table 4). AUCs ranged from 0.76-0.9 for ciprofloxacin, 0.69-0.82 for ceftriaxone, 0.66-0.77 for gentamicin, and 0.65-0.75 for trimethoprim-sulfamethoxazole. For ertapenem, we performed internal derivation for Dataset 1 and Dataset 3, and found apparent and optimism adjusted AUCs ranging from 0.74-0.98 and 0.7-0.98 respectively (Supplementary Table 2).

### ST Reference Database Approach (Lineage-based)

We calculated the AUCs and test characteristics for predicting antibiotic susceptibility for isolates in Dataset 2 with an *ST Reference Database*, using a variety of derivation datasets (Supplementary Table 5). AUCs ranged from 0.85-0.95 for ciprofloxacin, 0.67-0.85 for ceftriaxone, 0.73-0.83 for gentamicin, and 0.56-0.8 for trimethoprim-sulfamethoxazole.

### Best Genetic Match Reference Database Approach (Nearest neighbor)

We calculated the AUCs and test characteristics for predicting antibiotic susceptibility for isolates in Dataset 2 with a *Best Genetic Match Reference Database*, using a variety of derivation datasets (Supplementary Table 6). For all isolates, AUCs ranged from 0.83-0.92 for ciprofloxacin, 0.58-0.72 for ceftriaxone, 0.65-0.66 for gentamicin, and 0.53-0.63 for trimethoprim-sulfamethoxazole. For the top 25%’tile of best match datasets, AUCs ranged from 0.85-0.97 for ciprofloxacin, 0.57-0.88 for ceftriaxone, 0.76-0.86 for gentamicin, and 0.54-0.73 for trimethoprim-sulfamethoxazole. In any method based on comparing new samples with an existing database, it is important to investigate how large the database needs to be to permit accurate prediction. Thus, we evaluated the impact of varying reference database sizes on the performance of the genetic distance approach, with results shown in Figure S1.

### Summary of Test Characteristics Across Models

In Figures 2a-d, we summarize the post-test probabilities of susceptibility for: (1) positive model predictions indicating a likely susceptible isolate (PPV [positive predictive value]); and (2) for negative model predictions indicating a likely resistant isolate (1-NPV [Negative predictive value]). Here we see that the models that provide the best positive and negative predictive values are the *ST Reference Database*, the *Best Genetic Match Reference Database*, and combinations of the two (Supplementary Table 7). We also see that for most antibiotics and models, that the post-test probabilities for results indicating a susceptible result are sufficiently high (relative to syndromic thresholds) to support recommendation of therapy. Similarly, for most antibiotics and models the post-test probabilities of results indicating resistance is sufficiently low to support withholding therapy for a particular agent. Lastly, combined derivation datasets tend to have the most consistent model performance.

**Figure 2a.**
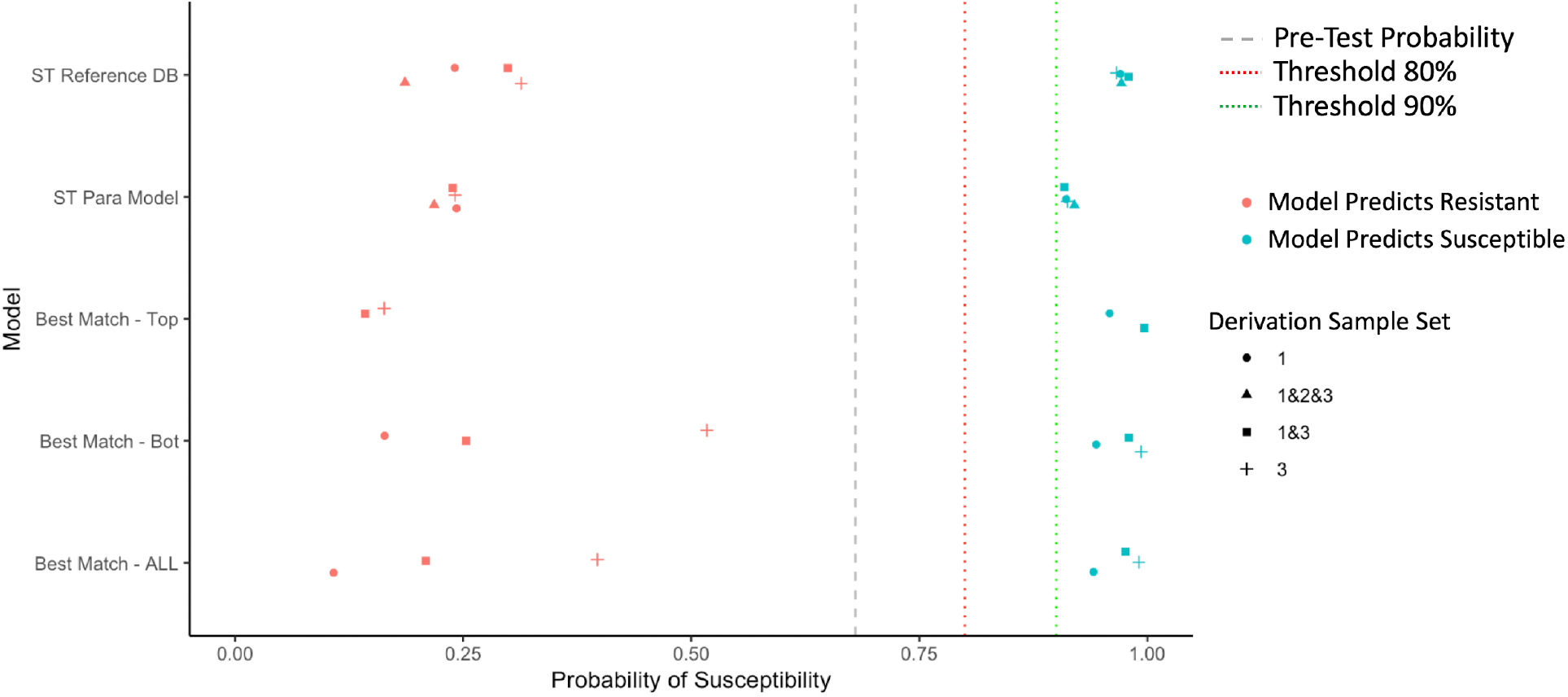
Selected post-test probabilities of trimethoprim-sulfamethoxazole susceptibility (in Dataset 2) based on Model Predictions of Resistant or Susceptible, by model type and derivation dataset.

**Figure 2b.**
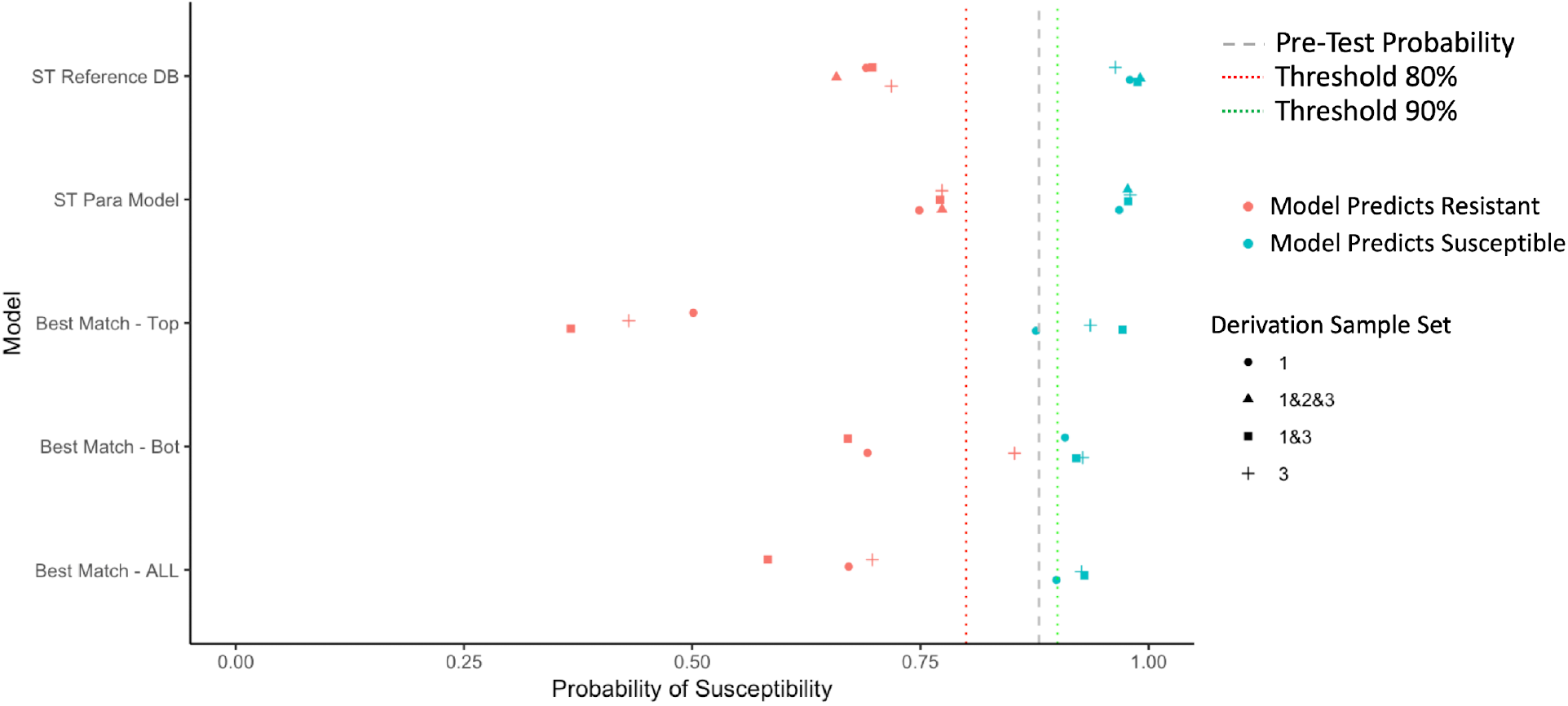
Selected post-test probabilities of gentamicin susceptibility (in Dataset 2) based on Model Predictions of Resistant or Susceptible, by model type and derivation dataset.

**Figure 2c.**
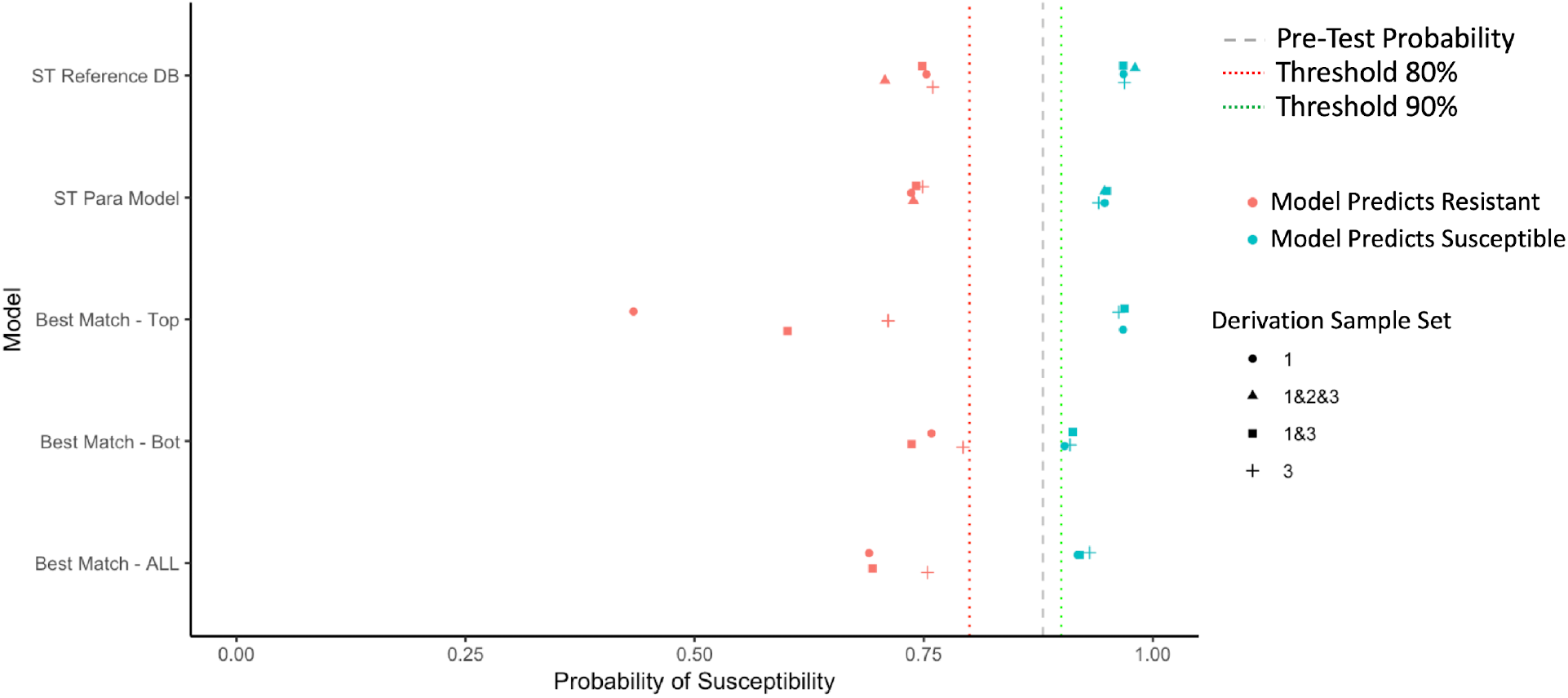
Selected post-test probabilities of ceftriaxone susceptibility (in Dataset 2) based on Model Predictions of Resistant or Susceptible, by model type and derivation dataset.

**Figure 2d.**
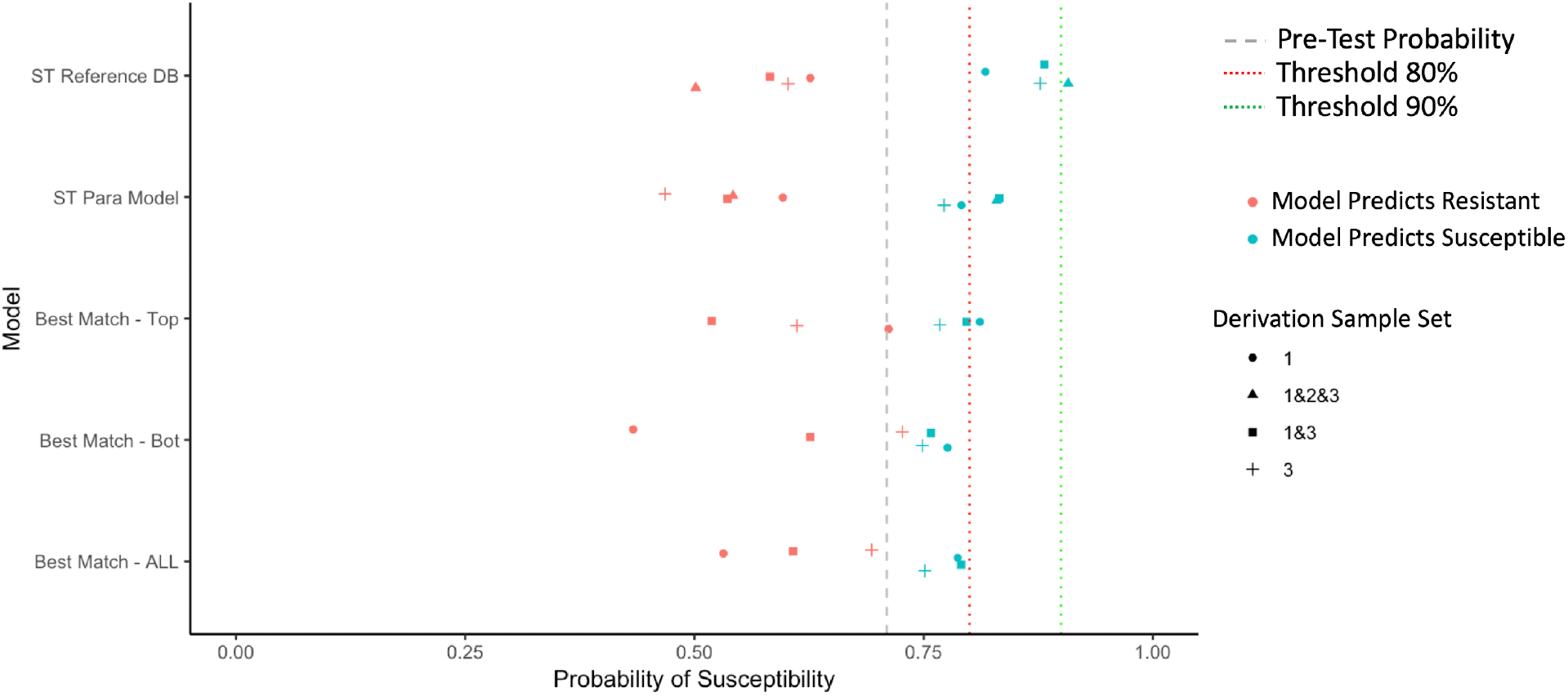
Selected post-test probabilities of ciprofloxacin susceptibility (in Dataset 2) based on Model Predictions of Resistant or Susceptible, by model type and derivation dataset. *The aim of the models is to shift green data-points to the right and red data points to the left.

### Sequential Antibiotic Decision-Making Based Upon a Prespecified Antibiotic Cascade

When using a sequential selection model favouring narrow spectrum antibiotics, adequate therapy was achieved for 89-98% of recommendations, with 47-70% of recommendations being a narrower spectrum agent with oral formulation (i.e. ciprofloxacin or trimethoprim-sulfamethoxazole). Spectrum scores for the narrow spectrum model cascade were consistently lower than those attained with the employment of typical empiric agents (Supplementary Table 7). Using an adequacy focused cascade, adequate therapy was achieved for 98-100% of recommendations, with 0-5% of recommendations being narrower spectrum agents with oral formulation options. Spectrum scores were generally high, but were lower than the most broad spectrum agent ertapenem.

To summarize, when using a narrow-spectrum focused cascade, the models yield excellent adequacy while enabling over ⅔ of recommendations to be narrow spectrum oral agents. When using an adequacy focused cascade they yield close to perfect adequacy but at the expense of using broader spectrum agents (Supplementary Table 7).

## Discussion

In this study, we demonstrate that we can predict the resistance phenotype of *E.coli* by rapidly determining the genetic relatedness of the infecting pathogen to a database of sequenced isolates with known resistance phenotypes. We show that these relationships can be used to generate post-test probabilities of susceptibility in excess of 0.8 or 0.9 which render antibiotics with a high prevalence of resistance to be empirically useful (e.g. ciprofloxacin or trimethoprim-sulfamethoxazole for urinary tract infection). In essence, adoption of this approach would modify the current stages of empiric therapy to introduce a new window that is informed by genetic relatedness, and information that is available in advance of standard phenotypic testing. This approach is a supplement to, not a replacement for, gold standard phenotype. The contribution of such a system could be to improve the quality of antimicrobial prescribing in the window between culture positivity and phenotypic testing results, by 1) reducing the expected time from culture positivity to adequate treatment and 2) reducing the duration of use of broad-spectrum agents to treat infections that could be adequately treated with a narrow-spectrum agent.

Looking at the distribution of STs across the datasets (Figure 1, Table 1) we see that Dataset 3 is enriched for ST131, and this is presumably due to the intentionally biased sampling approach toward MDR. However, it means that lineage and nearest neighbor approaches will be able to draw sample phenotype predictions from different datasets based on genetic proximity, not simply ones most closely related on temporal, geographic, or anatomic scales.

When predicting antibiotic resistance based on lineage, *ST Parametric Model* and *Cluster Parametric Model Approaches* (Supplementary Tables 3 and 4) provide reasonable discrimination for ciprofloxacin susceptibility (AUCs 0.76 - 0.91), in keeping with existing literature[15]. This emphasizes the strong association between lineage and ciprofloxacin susceptibility. By contrast there were only modest associations for the other antibiotic classes (AUCs 0.6-0.82). For all antibiotic classes, the use of combined derivation datasets (1 and 2 and 3 or 1 and 3) seemed to perform well most consistently, and this supports the use of an aggregated derivation dataset across time, geography, and anatomic location.

Using an *ST Reference Database* approach has the appeal of providing improved predictions for less common STs. When considering only those isolates with a matching ST in the reference database, the discrimination of this approach paralleled and sometimes exceeded that of *ST* and *Cluster Parametric Models* (Supplementary Table 5). The notable downside to this approach is the inability to provide predictions for sequence types outside of the reference database, though the proportion of these were small and decreased with increasing reference database size.

A *Best Genetic Match Reference Database Approach* (nearest neighbor) offers potential improvement over the *ST Reference Database Approach* (lineage), in that it might improve predictive performance for those classes that have a weaker association with specific lineage (e.g. ceftriaxone, gentamicin, trimethoprim-sulfamethoxazole). The genetic distance approach seemed to operate best under two circumstances: (1) when only the ‘top matches’ were considered and (2) when a combined derivation dataset was used. This ‘top match’ nearest neighbor approach is potentially implemented using a predefined threshold of Mash distance (or other genetic distance measure), but suffers from having a significant number of samples for which predictions may not be offered. Interestingly, the ‘top match’ approach can improve AUCs compared to other approaches for the antibiotics that are less strongly associated with phenotype (ceftriaxone, gentamicin, trimethoprim-sulfamethoxazole).

One important consideration with a genetic distance based model, is the consideration of how large the reference database should be. Using repeated sampling, we found that optimal performance could be achieved with relatively small derivation datasets, consisting of 100-200 samples. However, the necessary size will likely depend on the diversity of the population being evaluated, with more diverse populations requiring larger databases. This is reflected in the performance seen with combined derivation datasets.

There are limitations to our study. First, we were only able to consider the construction of reference databases confined to the geographic region of the province of Ontario. However, this region is geographically large and contains a population of over 14 million people. As such, our results still support construction of regional databases at least at this jurisdictional level, which is generalizable to many areas globally. Secondly, we did not evaluate the utility of this approach for other bacterial species, however *E.coli* is the most common Gram-negative pathogen in the hospital and community. Future work will aim to prospectively evaluate these techniques in a clinical setting using rapid sequencing approaches across geographic scales and additional pathogens. In summary, our results suggest that rapidly obtainable genomic information from clinical isolates can support intelligent choices that improve empiric antibiotic therapy, both by rescuing narrow spectrum agents for therapeutic use and by better selecting broader spectrum agents.

## Data Availability

Genomic data will be uploaded and available through the National Center for Biotechnology Information’s Sequence Read Archive.

## Acknowledgements

There are no additional acknowledgements.

